# Characterization of beat frequency artifacts in dual-device deep brain stimulation

**DOI:** 10.1101/2025.10.11.25337803

**Authors:** Nabeel Diab, Saipravallika Chamarthi, Kara Presbrey, Stephanie Cernera, Sameer Rajesh, Raphael Bechtold, Nisha Giridharan, Garrett Banks, Eric A. Storch, Doris D. Wang, Philip A. Starr, Wayne K. Goodman, Jeffrey A. Herron, Sameer A. Sheth, Nicole R. Provenza

## Abstract

**Objective:** Recording-capable deep brain stimulation systems enable chronic monitoring of neural activity concurrently with stimulation. DBS configurations with two implantable pulse generators (IPGs) are becoming more common to increase the number of sensing and stimulating channels. However, even when both IPGs are programmed to the same stimulation frequency, slight mismatches IPG clock rate can cause deviations in true stimulation frequency across IPGs. These timing discrepancies create high amplitude “beat frequency artifacts” (BFAs) at regular intervals, complicating the interpretation of neural data. This study aimed to characterize the occurrence and timing of BFAs in dual-IPG systems and determine their relationship to differences in stimulation frequency.

**Approach:** In one patient, we intentionally created a DBS frequency mismatch by stimulating at different frequencies across the two IPGs. In a cohort of 26 patients, we quantified BFA intervals in LFP recordings during which stimulation frequencies were set to identical values on both IPGs during both continuous and adaptive stimulation modes.

**Main Results:** We observed a predictable relationship between BFAs and stimulation frequency and found that BFAs occurred at a rate of *1/Δ frequency*, such that greater differences in DBS frequency resulted in more frequent artifacts. Even when both IPGs were set to the same stimulation frequency, BFAs occurred for all patients. During continuous DBS, artifact intervals ranged from 112 seconds to 30 minutes across the cohort of 26 patients. In a single patient, adaptive DBS (aDBS) mode shortened BFA intervals, suggesting that the transition to adaptive mode may influence the timing relationship between IPGs.

**Significance:** Recording neural activity during concurrent stimulation from two DBS devices leads to unique artifacts that may be caused by small discrepancies in effective stimulation frequency arising from independent device clocks. The interval between artifacts is predictable based on the DBS frequency mismatch, and may be exacerbated in certain stimulation modes, including aDBS.

## Introduction

With the commercial growth of recording-capable deep brain stimulation (DBS) devices,^1^ researchers have begun to use on-device neural records to inform therapeutic stimulation delivery.^2–10^ In select patients, offering two implantable pulse generators (IPG) may expand the therapeutic and scientific utility of these systems.^5,6,11–16^ First, dual-device implantation doubles the number of leads available for stimulation and local field potential (LFP) recording, enabling treatment of comorbid conditions that may require distinct stimulation targets. Second, utilizing a second device doubles the number of recording channels that can be distributed across recording and stimulating electrodes. This expanded neural sampling can improve characterization of pathological neural mechanisms as well as support more flexible adaptive stimulation algorithms.

Since each device operates independently of the other, stimulation pulses are delivered at a frequency dictated by an internal clock on each device. These clocks count time based on the oscillation frequency of an embedded quartz crystal. Since no two crystals are identical, no two clocks operate at exactly the same rate. This slight mismatch in internal clocks across IPGs causes a slight mismatch between identically programmed stimulation frequencies across two IPGs. As in any situation where two oscillators are producing different frequency inputs into a system, a lower frequency “beat” occurs with a frequency equal to the frequency difference between the two oscillators. We thus hypothesize that in patients with two DBS IPGs, the effect of small stimulation frequency differences would result in beat frequency artifacts (BFAs) in the neural data stream.

In 26 patients implanted with two DBS IPGs, we show that delivering stimulation from two devices produces BFAs in the recorded neural data. These BFAs are high amplitude deviations from biological activity that occur at regular intervals and contaminate recordings of neural activity.

Left unaddressed, BFAs hinder LFP analyses and the identification of neural biomarkers needed for optimization of neuromodulation therapy and adaptive DBS (aDBS) strategies.

## Methods

### Study Participants

26 patients at two institutions (Baylor College of Medicine [BCM] and University of California at San Francisco [UCSF]) were implanted with two Medtronic Summit RC + S devices with one IPG placed in each side of the patient’s chest. The local institutional review board at BCM (H-49155) and UCSF (18-24454 and 20-32847) approved all procedures. At BCM, we implanted five patients with treatment refractory obsessive-compulsive disorder (OCD) with bilateral DBS leads in the ventral capsule/ventral striatum (VC/VS) and bilateral electrocorticography (ECoG) strips over the orbitofrontal cortex (OFC) (NCT04806516). At UCSF, we implanted 21 patients with Parkinson’s Disease (PD) with bilateral DBS leads in either the subthalamic nucleus (STN) or globus pallidus internus (GPi), in addition to ECoG strips over the primary motor cortex (M1) (NCT03582891 and NCT04675398).

### Electrode and Stimulation Configuration

Each Medtronic Summit RC + S electrode contains contacts numbered 0-3. We performed monopolar stimulation bilaterally at one of the two middle contacts (contact 1 or 2) in either the VC/VS, STN, or GPi, across all patients. Of the four bipolar recording channels per IPG, two channels recorded activity from the stimulating DBS lead and the other two recorded from the ECoG strips. The two recording channels on the DBS lead flanked the stimulating contact to reduce stimulation artifact when recording. The two recording channels on each ECoG strip consisted of adjacent pairs (contact pairs 8-9, 10-11). We delivered stimulation in the continuous DBS (cDBS) mode both at home and in clinic except during experimental delivery of aDBS through the device’s onboard “Operative aDBS” mode in the clinic. Neural recordings all passed through an onboard 0.85 Hz high pass filter and 100 Hz low pass filter.

### Calculating BFAs

We define the frequency difference (Δf) across device 1 (f_1_) and device 2 (f_2_) with the following calculation:

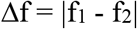

Where the frequency of BFAs is equal to Δf. Therefore, the interval between BFAs is inversely related to Δf in any given system.

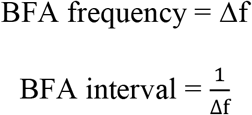

Hence, higher frequency BFAs have a shorter BFA interval and a greater Δf.

#### Experiment 1 - Frequency Modulation Testing

To test the impact of varying stimulation frequency across the two IPGs on observed BFA periodicity in a single patient, we modified the frequency of DBS for one OCD DBS patient (bilateral VC/VS electrodes and OFC ECoG strips) receiving therapeutic stimulation at 5.5 mA, 150.6 Hz, and 120 µs bilaterally. Without changing left hemisphere stimulation parameters, we decreased the right side stimulation frequency to 146.2 Hz while keeping right side stimulation amplitude and pulse width constant, which we expected would result in a BFA with a frequency of 4.4 Hz. After 217 seconds, we increased the right side stimulation frequency to 148.8 Hz, which we predicted would result in a BFA with a frequency of 1.8 Hz. We returned the right side stimulation frequency to 150.6 Hz after 300 seconds of total experiment time. Any BFA observed in this configuration would occur due to quartz-crystal variability with a very low (<0.1 Hz) Δf.

#### Experiment 2 - Unilateral DBS OFF Testing

To demonstrate that BFAs originate from stimulation interference across devices, we recorded LFPs in both devices while only one was stimulating at a time. Cessation of the artifact would confirm that the observed artifacts are BFAs rather than artifacts from another source. For all DBS off testing (n=3), we turned only one device off at a time. Since stimulation at 0 mA produces a stimulation artifact on recording channels,^8,17^ we switched the devices to DBS off rather than decreasing stimulation amplitude to 0 mA. In Figure 1B, both devices continued to stimulate at clinical settings before we turned off stimulation in the right device for 160 seconds. We then turned right side stimulation back on to clinical settings for another 230 seconds before turning the left side stimulation off for 150 seconds. We turned left side stimulation back on and maintained clinical stimulation parameters until the experiment ended.

**Figure 1.**
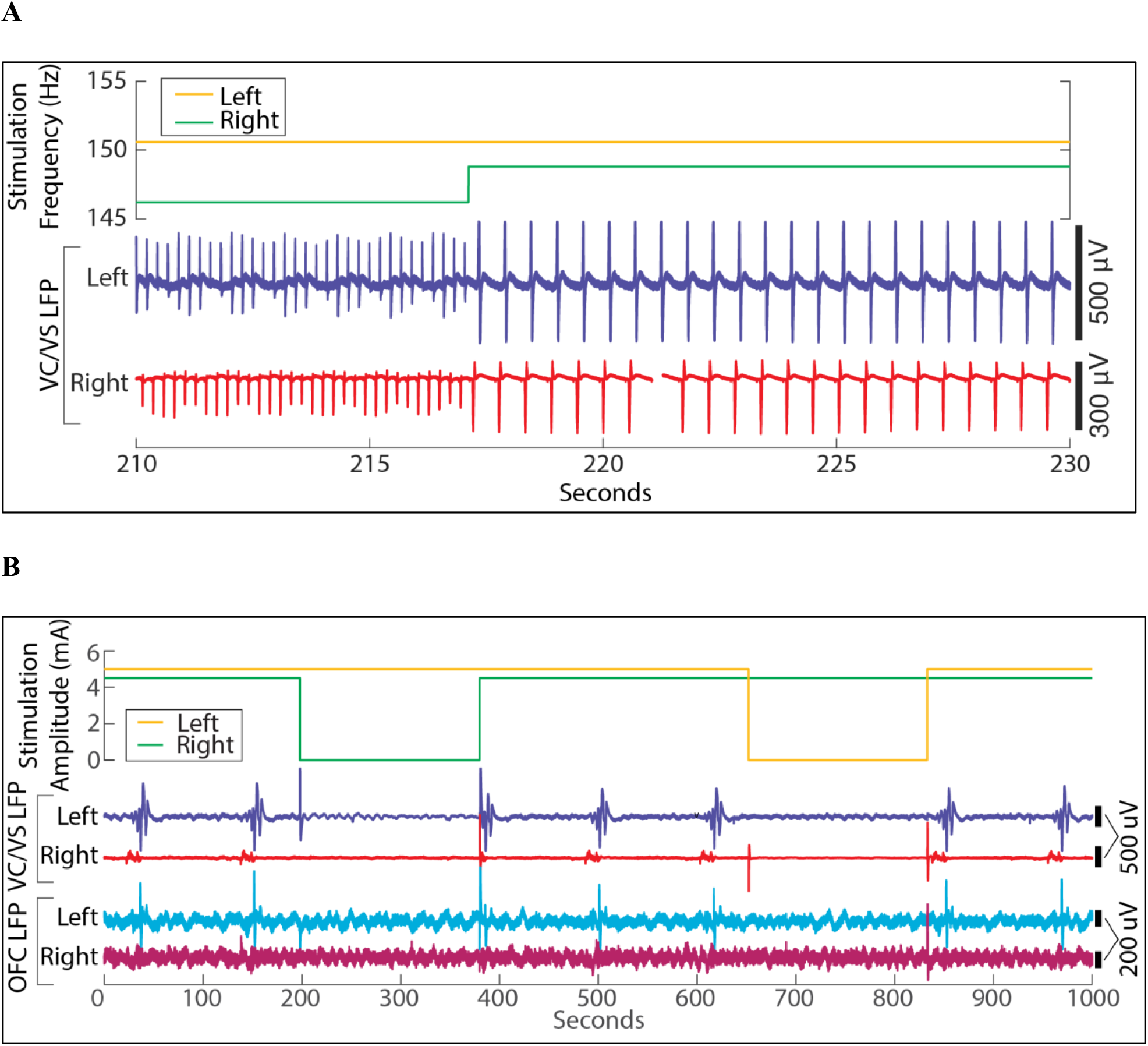
Beat frequency artifacts occur when devices are set to slightly different stimulation frequencies. (A) DBS frequency and VC/VS LFP over time. Left IPG stimulation is held at 150.6 Hz over the 20 second window. Right IPG stimulation frequency is 146.2 Hz until 217 seconds which produces a 4.4 Hz BFA equivalent to the difference in stimulation frequencies across devices. The BFA frequency drops to 1.8 Hz when right IPG stimulation frequency increases to 148.8 Hz (n=1). (B) DBS amplitude, VC/VS LFP, and OFC LFP over time. Beat frequency artifacts are only present when both devices are actively stimulating. Data from 1 of 3 patients tested with one device off is shown. BFAs contaminate neural signals on both stimulating leads in the ventral capsule/ventral striatum (VC/VS) and cortical electrode recording channels in the orbitofrontal cortex (OFC).

### Long-term Recordings

To evaluate the impact of BFAs on long-term recording and data collection across our entire patient cohort, we collected single LFP recordings exceeding 1000 seconds in length from either in-clinic behavioral tasks or passively at home from each patient across both institutions. We determined the presence of BFAs in each recording by identifying more than three stereotypical artifacts that occurred at exactly the same interval. After determining the presence of BFAs in each recording, we assessed across-patient differences.

We also visually identified “cap stack” artifacts within the recordings to distinguish them from study-relevant BFAs. These artifacts manifested as ∼200 ms fixed, narrow amplitude deviations that occurred once every 300 seconds. Notably, cap stack artifacts are unique to the Medtronic DBS devices and result from proprietary stimulation engine optimization processes that periodically ensure accurate pulse delivery as device battery levels fluctuate over time. As this process occurs independently within each device, these artifacts were not observed simultaneously across recordings from different devices.

#### Experiment 3 - Amplitude Ramping Testing

We used repeated step-like increments and decrements in stimulation amplitude, termed “ramping,” to further investigate the behavior of BFAs during IPG on-off behavior. We performed ramping with the DBS device set to the “Operative aDBS” mode to allow for manual control of stimulation amplitude. We completed this test in a single patient and switched from cDBS to aDBS before ramping amplitude down, turning DBS off, and then ramping amplitude back up.

## Results

### Beat frequency artifacts emerge when DBS frequencies differ across two IPGs

To generate a BFA in vivo, we intentionally mismatched DBS frequency temporarily across both IPGs as described in *Experiment 1*. We found that BFAs occur at predictable frequencies equal to the Δf between the two stimulating devices (Fig. 1A). First, we set the stimulation frequencies for the left and right hemispheres to 150.6 Hz and 146.2 Hz, respectively. At this Δf of 4.4 Hz, we found that BFAs occurred approximately 4.4 times per second. We then changed the stimulation frequency in the right hemisphere to 149.8 Hz, reducing the Δf to 1.8 Hz, and the BFAs occurred less frequently, at a rate of 1.8 times per second.

Even when two IPGs are set to the same stimulation frequencies, BFAs arise due to slight mismatches in the clock rate across each device. We found a BFA interval of 112 seconds when stimulating at 150.6 Hz on both IPGs in one patient (Fig. 1B). To demonstrate BFAs arise from two simultaneously stimulating devices, we turned off one IPG at a time, as described in

*Experiment 2*. When stimulation was turned off on either device, the BFA disappeared. Therefore, we concluded that this periodic signal was most likely a BFA.

### BFAs were present in long-term recordings across all 26 participants

Next, we sought to investigate the presence of BFAs in a large, multi-institutional cohort of patients with two recording-capable IPGs. Although both IPGs were programmed to deliver therapeutic stimulation at identical frequencies, we observed BFAs in all patients (n=26) at variable intervals (Fig. 2), reflecting slight differences in the effective stimulation frequencies of both devices. The shortest interval between BFAs observed was 112 seconds, and each of these BFAs lasted under 10 seconds. The longest interval between BFAs observed was over 30 minutes with each artifact lasting about 200 seconds. Within individual patients, BFA intervals were highly reproducible across recordings.

**Figure 2.**
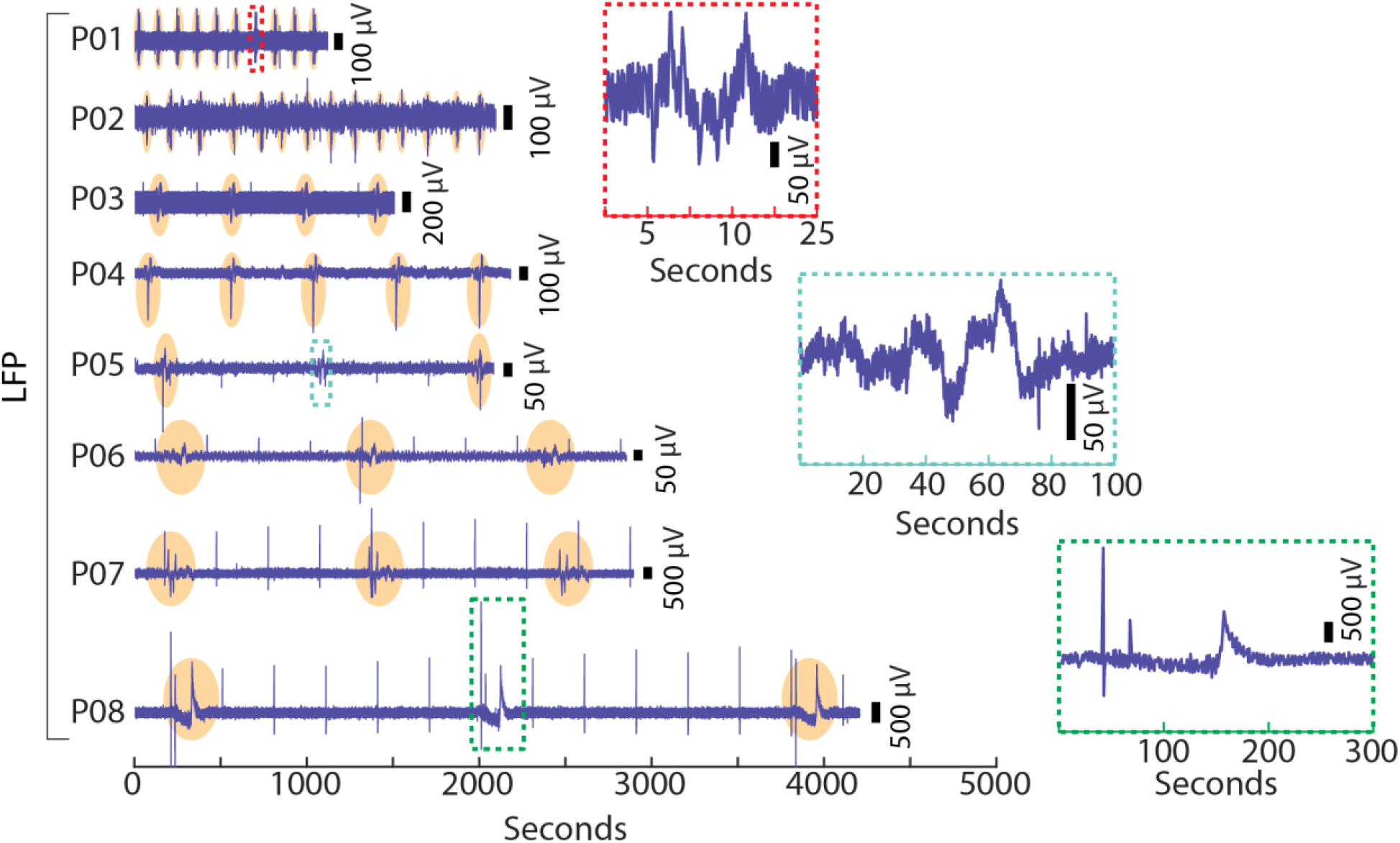
Longer intervals between beat frequency artifacts are associated with longer artifact durations. LFP recordings over time in a subset of 8 patients (P01-P08). Recordings were performed on DBS electrode recording channels except in P02 where a cortical channel was used. Patients are arranged by ascending BFA interval lengths from top to bottom and all recordings are during cDBS with equivalent stimulating frequencies. All BFAs are highlighted in orange except for callouts showing BFA examples in P01, P05, and P08 in red, blue, and green, respectively. Cap stack artifacts appear at 300 second intervals as sharp, narrow peaks in the LFP recordings of P03, and P05-P08 above.

### Adaptive DBS reduces the BFA interval

In a single patient, we investigated the impact of aDBS on BFA frequency and duration as described in *Experiment 3* (Fig. S1). To recreate the cDBS BFA shown in Figures 1 and 2, we conducted a baseline recording during which left and right devices were set to an amplitude of 5.5 and 5.7 mA respectively and frequency of 150.6 Hz bilaterally. During this baseline recording, we observed 3 BFAs at the expected interval of 112 seconds. After 12 minutes of baseline recording, we switched the right hemisphere device to “Operative aDBS” mode in order to automatically control state changes that guide DBS parameters. We decremented stimulation amplitude to 0 mA at a rate of 1 mA per minute. After reaching 0 mA, we turned DBS off for one minute, to avoid 0 mA artifact, before incrementing back to 6 mA at a rate of 3 mA per minute. While the right IPG was in aDBS mode, BFA intervals across both hemispheres decreased to ∼30 seconds during ramping of amplitude. This interval was not impacted by ramp rate. The amplitude of the BFA on the left was dependent on the stimulation amplitude of the right IPG. These higher-frequency BFAs took the place of the lower-frequency BFAs seen during baseline. As expected, BFAs disappeared in both hemispheres when one IPG was turned off.

**Figure S1.**
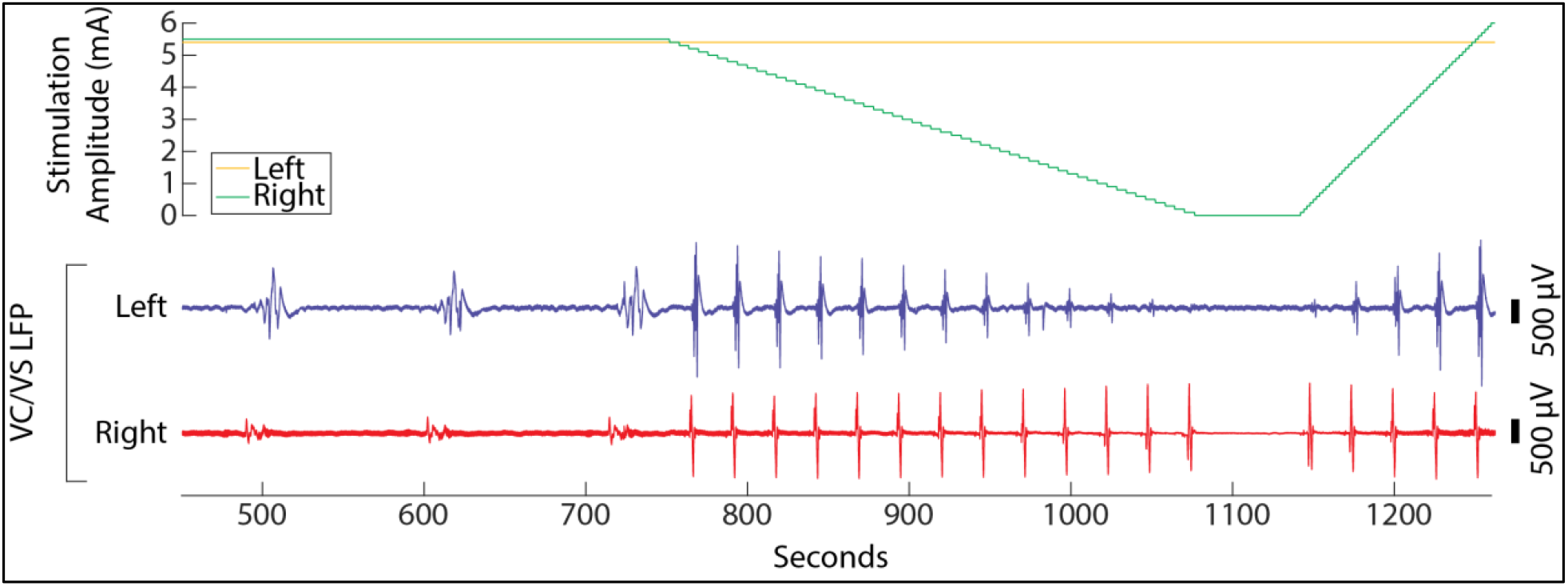
Decreased intervals between beat frequency artifacts in aDBS mode do not depend on ramp rate. After switching the right IPG from cDBS to aDBS and decrementing stimulation amplitude at 750 seconds, BFA intervals decrease from the 112 seconds during cDBS to 26.5 seconds and the artifact shortens in duration.

## Discussion

We demonstrate the presence of BFAs across all patients investigated (n=26) with two IPGs. These artifacts are unavoidable due to cross device interference when stimulating simultaneously. Each IPG relies on an internal clock derived from the oscillation frequency of an embedded quartz crystal. Due to minute differences, no two quartz crystals oscillate at exactly the same rate. Consequently, the duration of one second differs slightly across devices. When two devices are each set to a clinical stimulation frequency of 150.6 Hz, their actual stimulation frequencies will not match exactly. To the degree of precision of each IPG’s internal timestamp, these differences are negligible, but they are evidenced by reproducible and predictable BFAs. As the phase relationship between the two stimulation pulse trains gradually drifts, periods of constructive interference appear as large transient artifacts. Slower drift rates produce less frequent but longer-lasting artifacts, explaining the relationship between BFA interval and duration observed in Figure 2.

Because BFAs can persist for seconds to minutes, they may bias commonly used biomarker estimates such as band power, spectral features, or adaptive control signals derived from LFP recordings. When switching to aDBS, the IPG stimulation engine updates more frequently to adjust for frequent stimulation changes which could explain the decreased BFA interval seen in Fig. S1. To accommodate BFAs in future aDBS algorithms, it may be possible to ignore the high amplitude deviations of BFAs with the strategic implementation of onset times.^5^ Onset times are a Summit-defined embedded algorithm parameter which defines a minimum amount of time an adaptive state change threshold must be crossed before the stimulation change is implemented. Onset times longer than the BFA duration will allow aDBS algorithms to respond only to biological signals. This approach does however limit the responsiveness of aDBS algorithms.

Importantly, future work is required to characterize the impact of BFA artifacts onboard the commercial Medtronic Percept aDBS system.^18^ The impact of the BFAs on aDBS performance should be taken into consideration when deciding to leverage a dual-device approach, and BFAs specific to each patient should be characterized before analysis and adaptive algorithm development. Most patients we investigated had at least one recording channel with a very weak BFA, characterized by minimal amplitude deviation that could be chosen for aDBS implementation. Identifying which channels are least affected for each patient and adapting identified biomarker calculations accordingly can help researchers avoid the lengthening onset times while minimizing the presence of BFAs on aDBS algorithm inputs.

## Conclusion

This study establishes BFAs as a consistent and predictable phenomenon arising from simultaneous stimulation by dual IPGs in DBS therapy. Through controlled mismatches in stimulation frequency and long-term recordings across a multi-institutional patient cohort, we demonstrate that BFAs are an inherent consequence of minute discrepancies in internal clock rates between devices despite identical programming. We also show that aDBS modulates BFA characteristics, as ramping stimulation amplitudes led to shorter BFA intervals.

Given the dynamic nature of BFAs and their risk for interference with biomarker detection and adaptive algorithm performance, it is important to consider how BFAs may impact aDBS strategies implemented in the dual-device configuration. Ultimately, understanding and managing BFAs is necessary for implementing adaptive neuromodulation in patients with dual IPG configurations.

## Data Availability

All data produced in the present study are available upon reasonable request to the authors.

## Acknowledgements

The research was supported by the National Institutes of Health (NIH) NINDS BRAIN Initiative via contracts UH3NS100549 (S.A.S., W.K.G., N.R.P., J.A.H.) and UH3NS100544 (P.A.S.), NIH NINDS via contract R01NS130183 (D.D.W.), the McNair Foundation (N.R.P., S.A.S), Burroughs-Wellcome Trust CAMS Award (D.D.W), Chen Scholar (D.D.W.), and Michael J Fox Foundation (D.D.W.). Summit RC + S devices were donated by Medtronic as part of the BRAIN Initiative Public-Private Partnership Program. We thank Scott Stanslaski for providing technical expertise regarding the Medtronic DBS system. This work relied heavily on the community expertise and resources made available by the Open Mind Consortium (https://openmind-consortium.github.io/).

## Conflicts of Interest

W.K.G: Received donated devices from Medtronic. Consulting agreements with Biohaven Pharmaceuticals. S.A.S.: Consulting agreements with Boston Scientific, NeuroPace, Abbott, Koh Young, and Zimmer Biomet. Co-founder of Motif Neurotech. P.A.S.: consulting for InBrain Neuroelectronics Inc. and has grant funding (to support fellowships) from Medtronic Inc. and Boston Scientific Inc. D.D.W. consulting for Medtronic Inc., Boston Scientific Inc., and Iota Biosciences. Dr. Storch reports receiving research funding to his institution from the Ream Foundation, International OCD Foundation, and NIH. He was a consultant for Brainsway and Biohaven Pharmaceuticals in the past 24 months. He owns stock less than $5000 in NView (for distribution of the Y-BOCS and CY-BOCS). He receives book royalties from Elsevier, Wiley, Oxford, American Psychological Association, Guildford, Springer, Routledge, and Jessica Kingsley.

## Notes

### Author Declarations

The local institutional review board at Baylor College of Medicine (H-49155) and University of California at San Francisco (18-24454 and 20-32847) gave ethical approval for all procedures.

### Summary of Updates

The abstract and author list were updated with clarified methodology.

